# Use of a systems engineering framework to assess perceptions and practices about antimicrobial resistance of workers on large dairy farms in Wisconsin

**DOI:** 10.1101/2020.10.28.20221747

**Authors:** Ashley E. Kates, Mary Jo Knobloch, Ali Konkel, Amanda Young, Andrew Steinberger, John Shutske, Pamela L. Ruegg, Ajay K. Sethi, Tony Goldberg, Juliana Leite de Campos, Garret Suen, Nasia Safdar

**Affiliations:** University of Wisconsin-Madison, Madison, Wisconsin, USA; William S. Middleton Veterans Affairs Medical Center, Madison, Wisconsin, USA; Michigan State University, East Lansing, Michigan, USA

## Abstract

We studied farmworker practices potentially contributing to transmission of bacteria and antimicrobial resistant genes (ARGs) among animals and farm workers to identify human behavioral interventions to reduce exposure risk. Ten focus groups were conducted on eight farms to explore potentially high-risk practices and farmworker knowledge and experiences with antimicrobial use and resistance using the Systems Engineering in Patient Safety (SEIPS) framework. Farmworkers were asked to describe common tasks and the policies guiding these practices. We found workers demonstrated knowledge of the role of antibiotic stewardship in preventing the spread of ARGs. Knowledge of various forms of personal protective equipment was higher for workers who commonly reported glove-use. Knowledge regarding the importance of reducing ARG transmission varied but was greater than previously reported. Programs to reduce ARG spread on dairy farms should focus on proper hand hygiene and personal protective equipment use but at the level of knowledge, beliefs, and practices.

## Introduction

Antimicrobials are essential to both human and animal health (1). However, antibiotics are frequently overused, which is a major driver of antibiotic resistance. Antibiotic use in agriculture is concerning due to the risk of transmission of antimicrobial resistance genes (ARGs) to people (2). Usage of antimicrobials on dairy farms is a potential risk to human health by increasing the risk of exposure via foodstuffs as well as potentially driving selection of ARGs, although direct evidence is scant (3). However, for ARGs to “escape” from farms into the human population, critical events would have to occur involving human infection and transport. In particular, workers on dairy farms are in frequent close contact with cattle and cattle manure and may be at high risk of encountering potentially resistant pathogens (4). These workers may act as “entry points” for ARGs into the human population with the potential to travel beyond the dairy farm. To date, there is limited knowledge of how dairy farm worker health and safety practices and behaviors impact the transmission of ARGs within and beyond the farm. A wide range of activities and tasks take place on a dairy farm, and all pose varying levels of ARG exposure risk to workers (4, 5). For example, working with sick animals or handling manure may pose a greater risk for encountering enteric pathogens whereas working in the milking parlor with healthy cows may pose a lower risk. Largely missing from such studies, however, are assessments of the knowledge base of dairy workers and how this affects risk.

Here, we undertook a cross-sectional study to develop a better understanding of how dairy farm worker perceptions and routine practices may be associated with potential exposure to ARGs. Additionally, we aimed to identify potentially modifiable behaviors that could be targeted in future interventions to reduce exposure risks. We hypothesize that a potential lack of awareness may be a root cause of risky behaviors, policies, or practices (both at the worker and organization levels), which in turn may put workers at risk. We used a human factors and systems engineering framework which allows for a comprehensive assessment of the activities occurring on a farm.

## Methods

We conducted focus groups with dairy farm workers to understand the human and system level factors related to ARG transmission on farms and into the greater community. Focus groups were chosen over individual interviews as they allow for the generation of multiple perspectives in an interactive setting (6). To do this, we adapted the Systems Engineering Initiative for Patient Safety (SEIPS) model (7) for an agricultural setting. SEIPS highlights the interplay between work system elements (organization, environment, tasks, tools/technology) and people. The SEIPS model allows researchers to identify known barriers and facilitators to organizational outcomes which can contribute to the development of well-informed and targeted interventions to reduce exposure and other risk factors. Although SEIPS has been extensively used to examine healthcare work systems (8, 9), to our knowledge, the SEIPS model has not previously been used to examine agricultural work systems nor has it been used to specifically examine dairy operations. Figure 1 illustrates how the SEIPS model was adapted for the use on dairy farms in this study.

**Figure 1.**
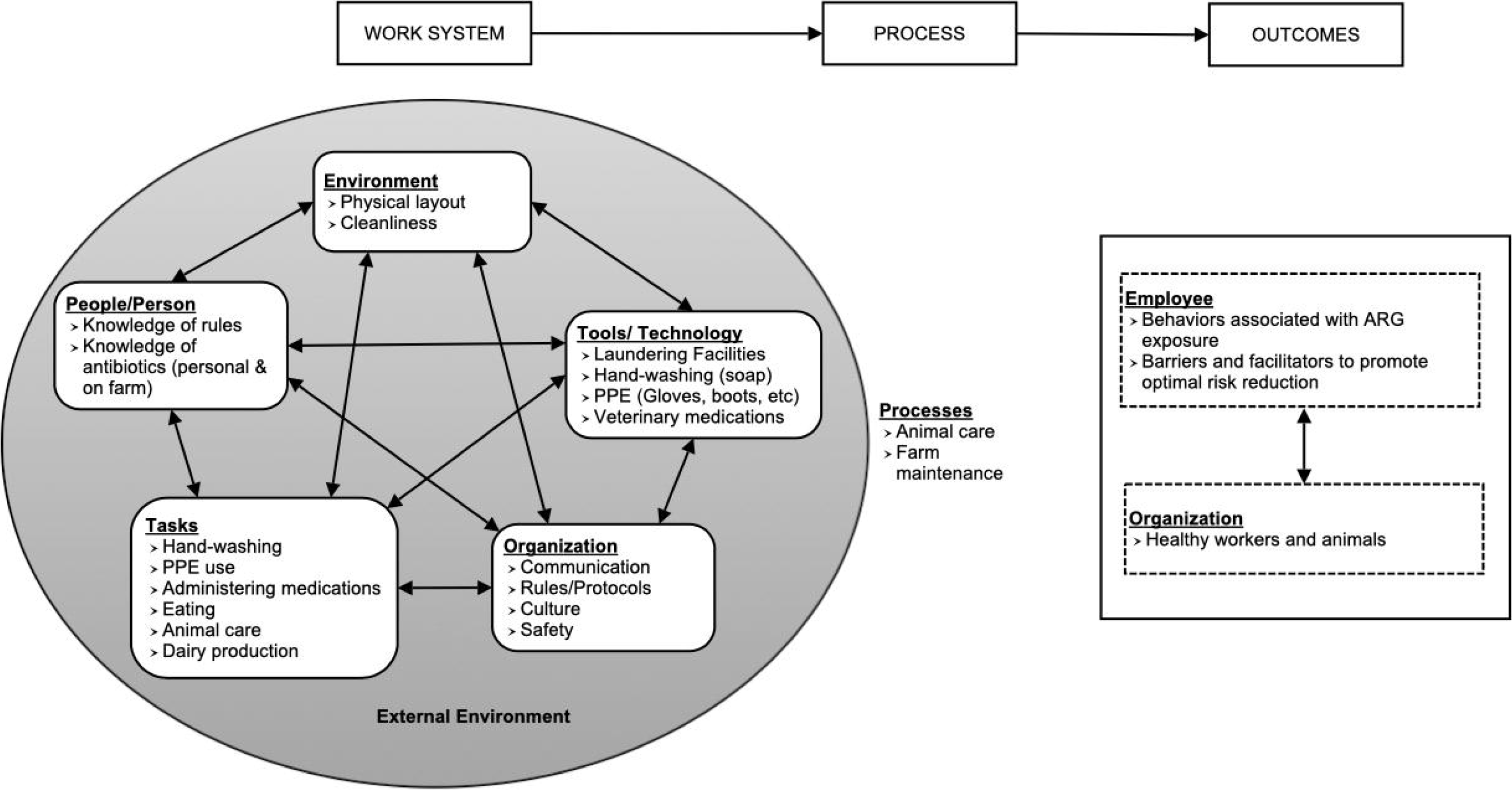
Systems Engineering in Patient Safety (SEIPS) model adapted for use on dairy farms. Headers represent the original SEIPS categories and bullet points represent how these categories related to dairy operations.

In this study, we explored daily routine practices, knowledge and experiences among farmworkers – obtaining a snapshot of activities related to antibiotic use\age on eight farms in Wisconsin selected to represent a range of antibiotic use in cattle. Farmworkers were asked to describe common tasks and work routines including hand hygiene, laundry and eating practices, use of personal protective equipment (PPE) and communication with managers in the context of farm guidelines and policies.

### Study population and recruitment

As part of a related study, we collected antimicrobial use data from 40 large dairy farms in Wisconsin and ranked farms based on daily doses of antibiotics per 1000 cow-days used (10). From this data, four low use farms and four high use farms were enrolled. Figure 2 provides an overview of study enrollment. Eligible farm owners were sent a letter inviting them to participate. When farms agreed to participate, owners were provided with study information posters (in English and Spanish) to be displayed in common areas. These posters alerted workers to the purpose of the research and the potential for researchers to visit the farm. Workers provided verbal informed consent and were compensated $25 for participation. All study documents and activities were approved by the University of Wisconsin-Madison Institutional Review Board (application ID: 2017-1333) prior to the start of research.

**Figure 2.**
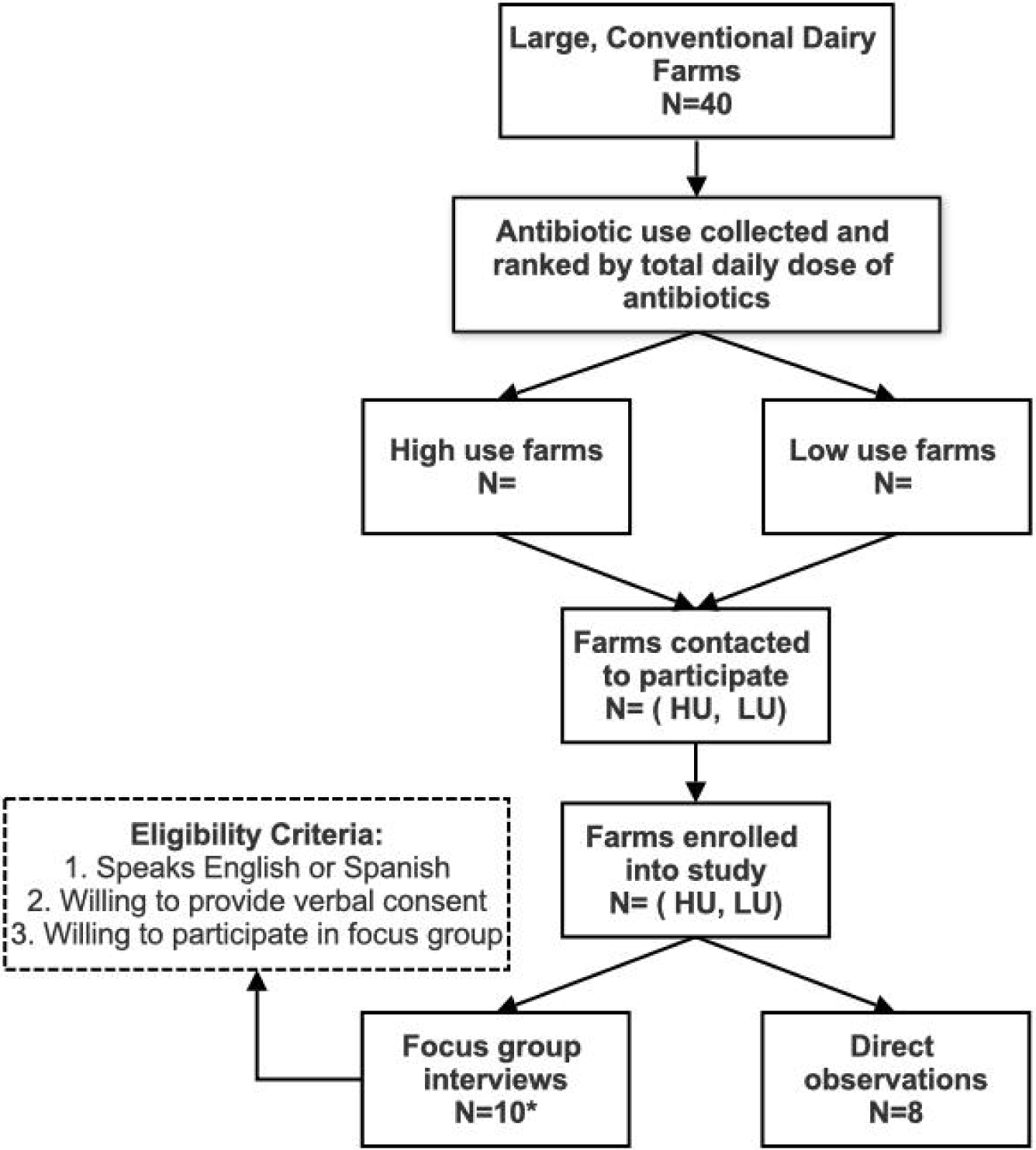
Flowchart of study enrollment. HU=high use farms; LU=Low use farms. *Two separate focus groups were conducted on two of the farms.

Focus group semi-structured interviews were conducted in both English and Spanish by trained researchers. The focus group question guide was developed by the research team according to the adapted SEIPS model. When possible, a representative sample of workers with a range of duties on the farm (calf care, sick animal care, maternity care, milking, farm maintenance, etc.) were invited to participate in the study. Depending on the size of the farm, workers may have been responsible for multiple types of tasks. Initial groups (n= 2) included both English and Spanish speakers. In subsequent groups (n=8), English and Spanish speakers were divided to allow for a more fluid discussion. Focus group discussions took place on the farm and farm managers/owners were asked to avoid the area where focus groups were conducted in order to allow workers to speak freely about their work life.

### Direct observations

On the same day, but prior to the focus group interviews, a researcher familiar with dairy farm practices conducted direct observations of defined animal management practices following a checklist developed by the research team (Appendix figure 1). The goal of this observation was to better understand the workflow on the farm related to infection prevention and to inform the assessment of barriers and facilitators identified during focus group interviews.

### Data analysis

Focus groups discussions were recorded, transcribed, and translated. Transcription and translation services were provided by Premium Business Services (Madison, WI). Dedoose v.8.0.35 was used to organize the data. We employed an iterative process to create the code book. Three researchers examined the same section of one transcript and compared and agreed upon codes and definitions. This process was used two additional times to adjust codes and definitions. Each element of the SEIPS model was considered for each section of the transcript and was discussed as a team as part of the first level of coding. Sub codes and definitions were identified for each element and agreed upon by researchers (second level of coding). Remaining transcripts were then coded by two researchers separately, and a third researcher with agricultural experience and qualitative expertise coded all transcripts identifying barriers and facilitators (third level of coding).

## Results

We conducted 10 focus groups across the eight farms enrolled into the study between December 2018 and October 2019. Observations of the facilities, equipment, PPE and worker behaviors were conducted on all eight farms prior to focus group interviews. A total of 60 farm workers participated in focus groups.

Table 1 lists SEIPS elements and sub-elements identified from the transcripts during the analysis as well as the number of times an element was coded as either a facilitator or a barrier to maintaining a healthy and safe work environment. Overall, facilitators (n=1041) were identified more often than barriers (n=307) across the farms. Representative quotations from each SEIPS element can be found in Table 2.

**Table 1.**
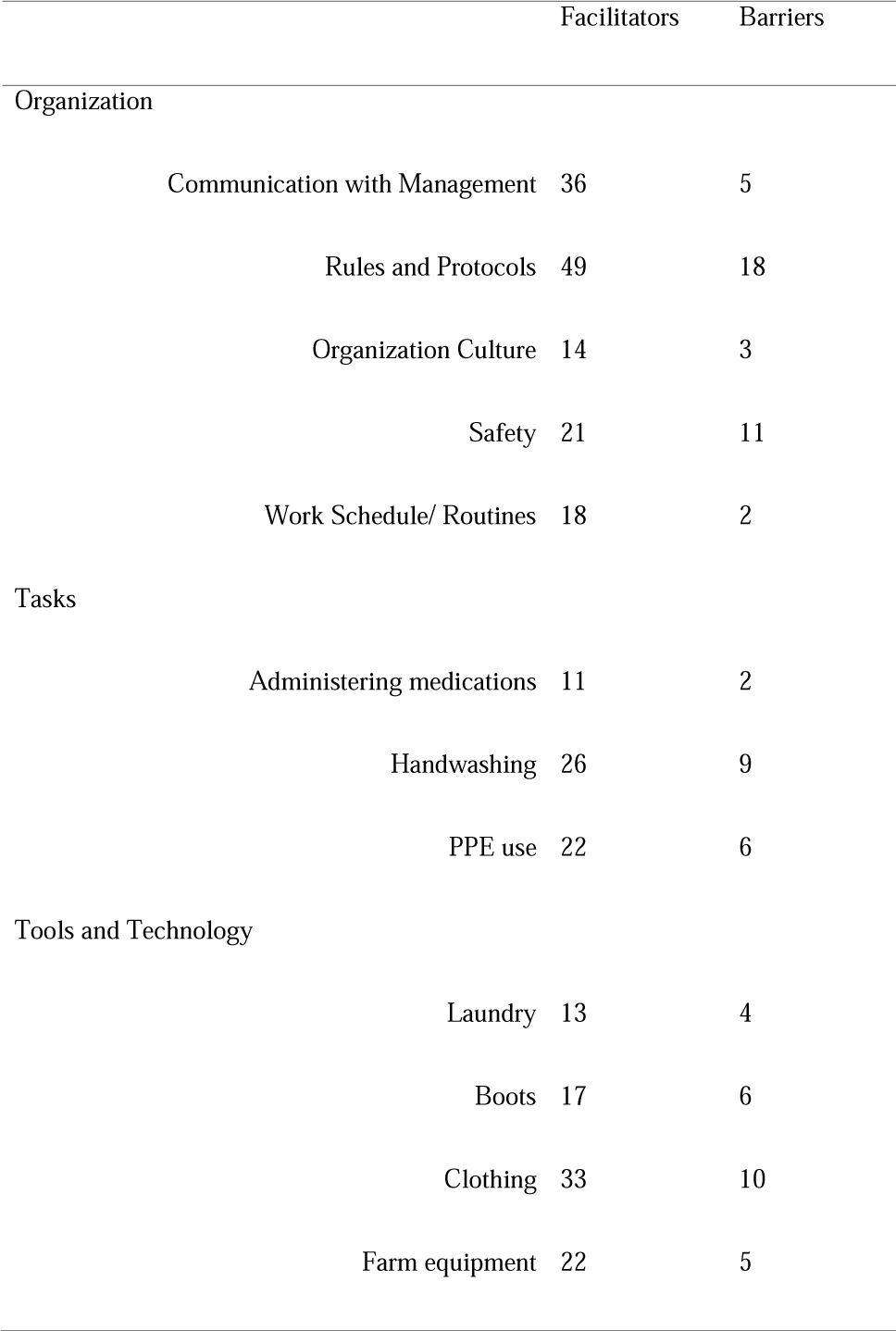
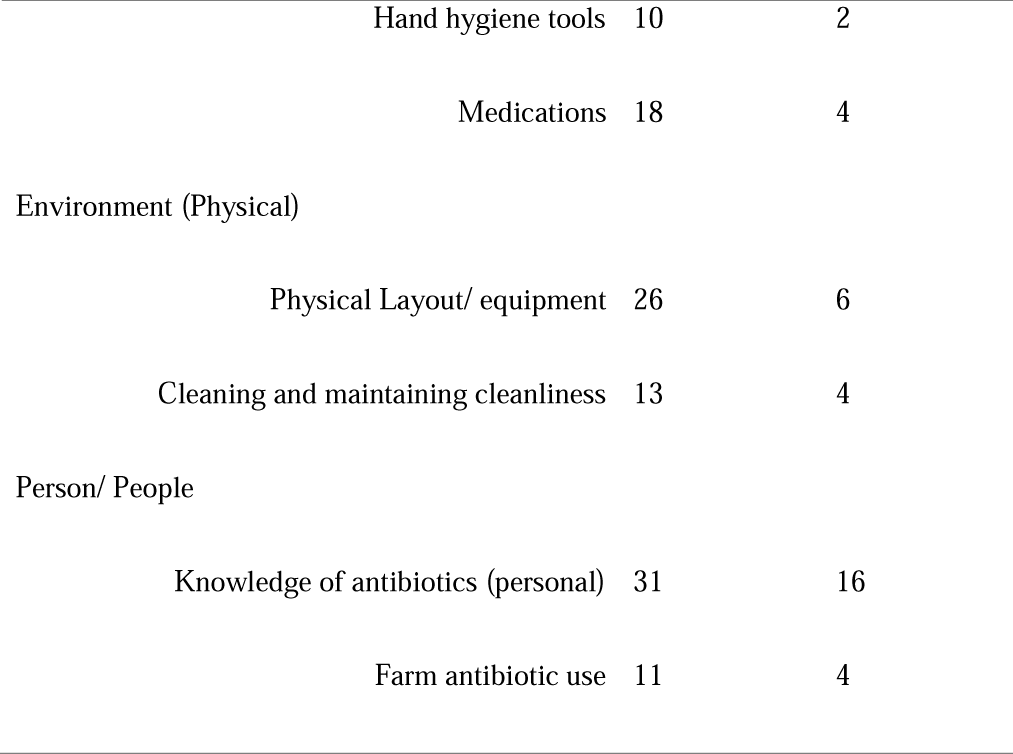
Number of barriers and facilitators associated with each SEIPS element.

|  | Facilitators | Barriers |
| --- | --- | --- |
| Organization |  |  |
| Communication with Management | 36 | 5 |
| Rules and Protocols | 49 | 18 |
| Organization Culture | 14 | 3 |
| Safety | 21 | 11 |
| Work Schedule/ Routines | 18 | 2 |
| Tasks |  |  |
| Administering medications | 11 | 2 |
| Handwashing | 26 | 9 |
| PPE use | 22 | 6 |
| Tools and Technology |  |  |
| Laundry | 13 | 4 |
| Boots | 17 | 6 |
| Clothing | 33 | 10 |
| Farm equipment | 22 | 5 |

|  |  |  |  |
| --- | --- | --- | --- |
|  | Hand hygiene tools | 10 | 2 |
|  | Medications | 18 | 4 |
| Environment (Physical) |  |  |  |
|  | Physical Layout/ equipment | 26 | 6 |
|  | Cleaning and maintaining cleanliness | 13 | 4 |
| Person/ People |  |  |  |
|  | Knowledge of antibiotics (personal) | 31 | 16 |
|  | Farm antibiotic use | 11 | 4 |

**Table 2.**
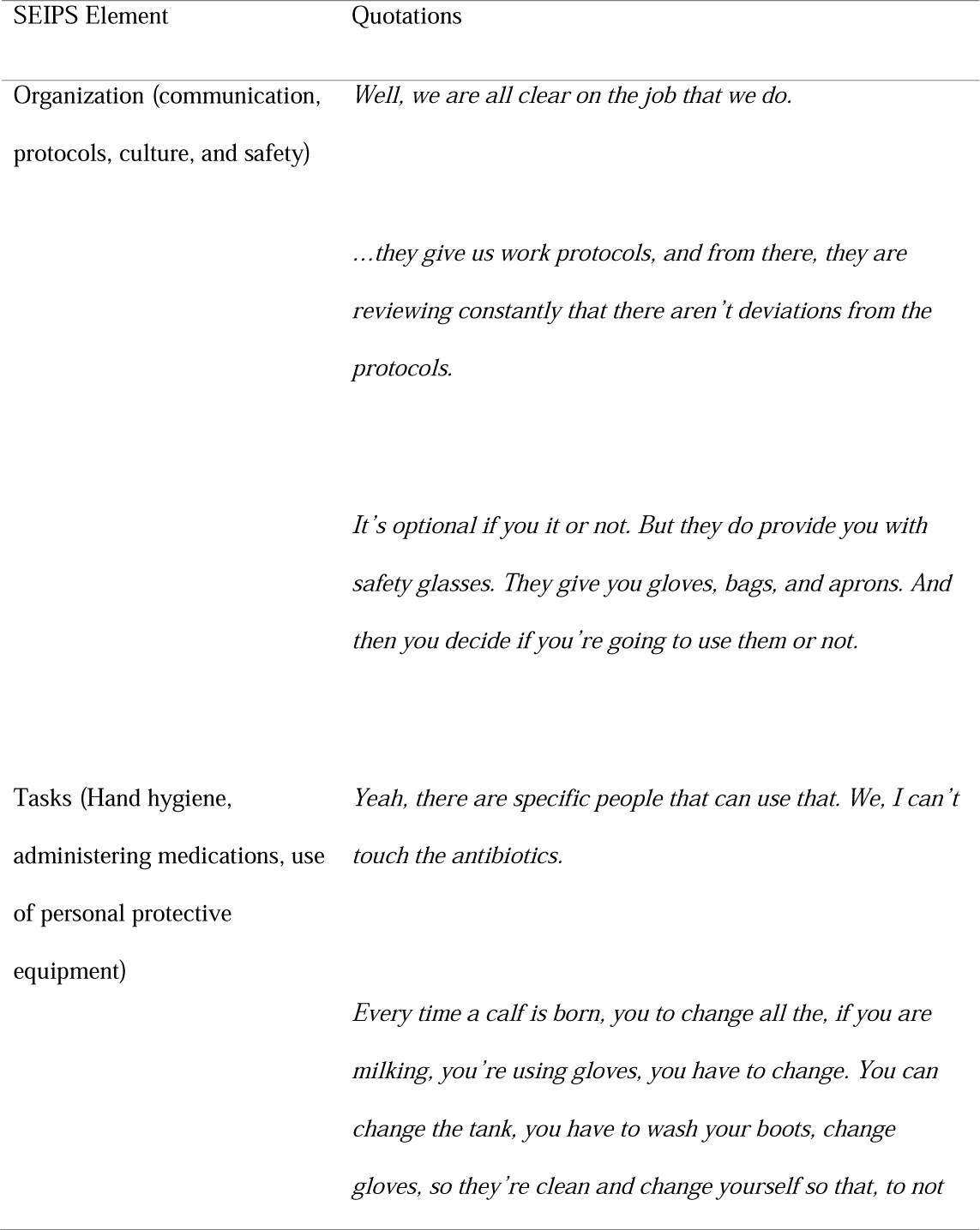
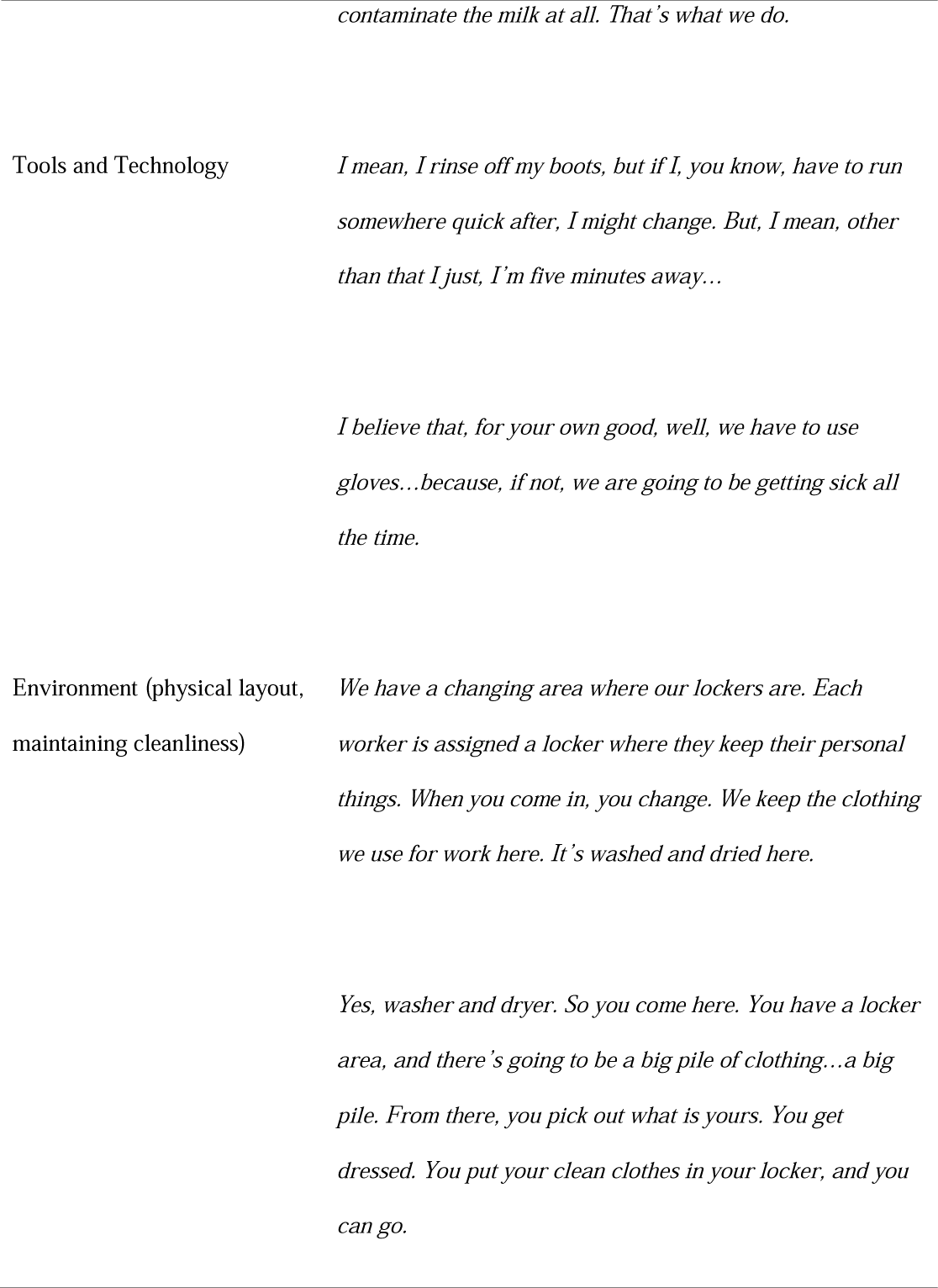
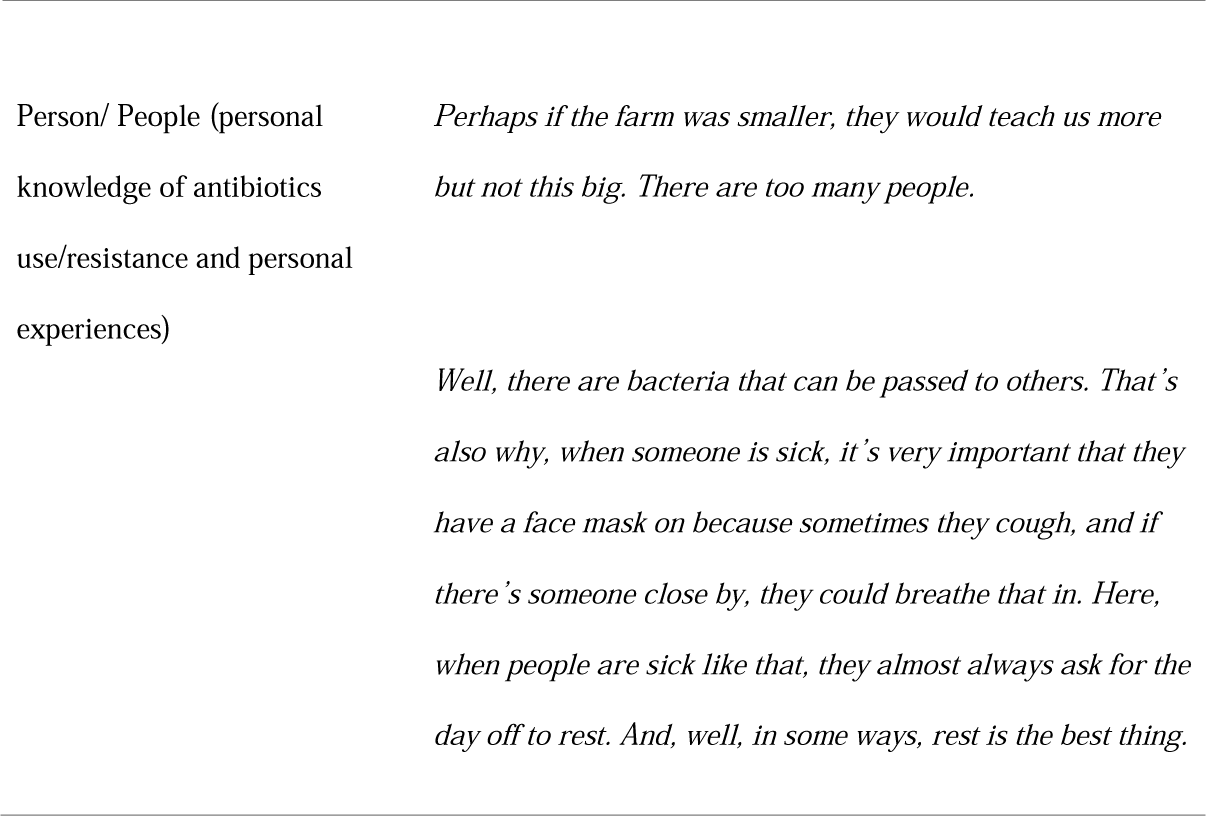
Representative quotes associated with each SEIPS element SEIPS Element Quotations.

### Organization

Many workers reported having knowledge of the farm’s protocols related to PPE use, safety, and administering antibiotics, although knowledge of rules or protocols was often identified often as a barrier. Workers reported having good relationships with farm management and their superiors and reported few issues in communicating with management, even in instances where language barriers existed.

> *“For that reason, we have restrictions because it could affect a lot. If we send milk to the tank that is contaminated with antibiotics, it’s a huge problem. So, no, it is very clear to us that we don’t give antibiotics, and that we keep them controlled. So only authorized people can do it”*

### Tasks

The culture around eating (meals and snacks eaten by workers) varied across farms. On some farms workers reported always eating only in the breakroom while on other farms, workers stated they ate wherever was most convenient or while performing their work duties, such as eating in the milking parlor or while driving around on the farm:

> *“We always eat in there. When I was on day crew, occasionally, they would, when they’re doing the expansions, they would have people in here, and they would be talking. So sometimes we would eat upstairs, or we would go into this room or that room”*

While six of the eight (75%) farms provided a clean place for preparing and eating food, the observer noted food wrappers and drink containers in other zones on four (50%) of the farms (Table 3).

**Table 3.**
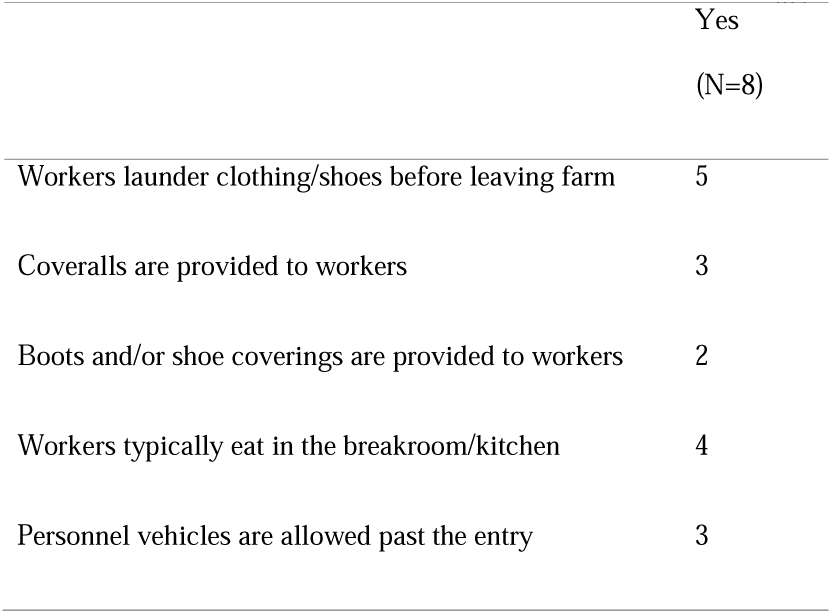
Observations of general biosafety practices on the farms.

Many workers demonstrated an understanding of the importance minimizing the spread of infectious organisms on the farm. Workers told us their farms had policies (either written or verbal) related to working with sick animals.

> *“They’ve gotten a bit more strict on the sick cattle and on the calving, when a cow is calving. They like it when you, they tell us that we have to keep our boots clean if we deal with a sick cow, and then we go in by a calf, we have to make sure that it is clean, because we don’t want to spread it*.*”*

### Tools and Technology

Worker perceptions on the tools and technology related to AMU on farms were variable. Use and availability of personal protective equipment (PPE) and availability of PPE was inconsistent across farms. While some farms provided coveralls and/or boots for workers to wear, others expected the worker to provide these items themselves. Some workers reported cleaning their boots before getting into personal vehicles or leaving for the day, while others said boot cleaning was not something they often did. Similarly, on some farms it seemed normal for workers to change out of their work clothes before leaving for the day and to wash them on the farm using the farms dedicated laundry equipment, while on other farms workers tended to wear their work clothes home. During the observations, the observer noted workers laundering work clothes on the farm and washing or changing boots before leaving on five farms (62.5%).

Many workers felt PPE use was important on the farm. When asked about handwashing practices, one worker stated:

> *“Yes, every time a calf is born, you have to change all the, if you are milking, you’re using gloves, you have to change. You change the [milk] tank, you have to wash your boots, change gloves, so they’re clean, and change yourself so that, to not contaminate the milk at all. That’s what we do*”.

Glove use was supported by observations where we identified 100% of workers using gloves in the milking parlor; most workers identified this practice as mandatory on their farm. On seven (87.5%) of the farms, workers were observed wearing PPE in the calf housing zones. Hand hygiene stations were available and contained appropriate materials in 100% of the bathroom and breakroom facilities in most high-risk areas. However, only three (37.5%) of the farms had a hand hygiene station in the hospital/isolation pen, although five (62.5%) provided easily accessible gloves in this zone. Easily accessible gloves were noticeably available in six (75%) milking parlors. Hand hygiene stations were only available in three (37.5%) of milking parlors (Table 4). Workers reported changing gloves most frequently after coming in contact with a sick and/or mastitis infected cow, eating, drinking, when gloves rip, when changing the line to the milk tank, and after going to the bathroom. Workers also reported they did not think there were necessarily specific rules around hand hygiene, but it was important to use common sense. Boot washing stations (presence of a hose at a minimum) were inconsistently available across farms, and not available in most zones. Calf housing was the exception to this with six (75%) of the farms having some type of boot washing station available near the entry.

**Table 4.**
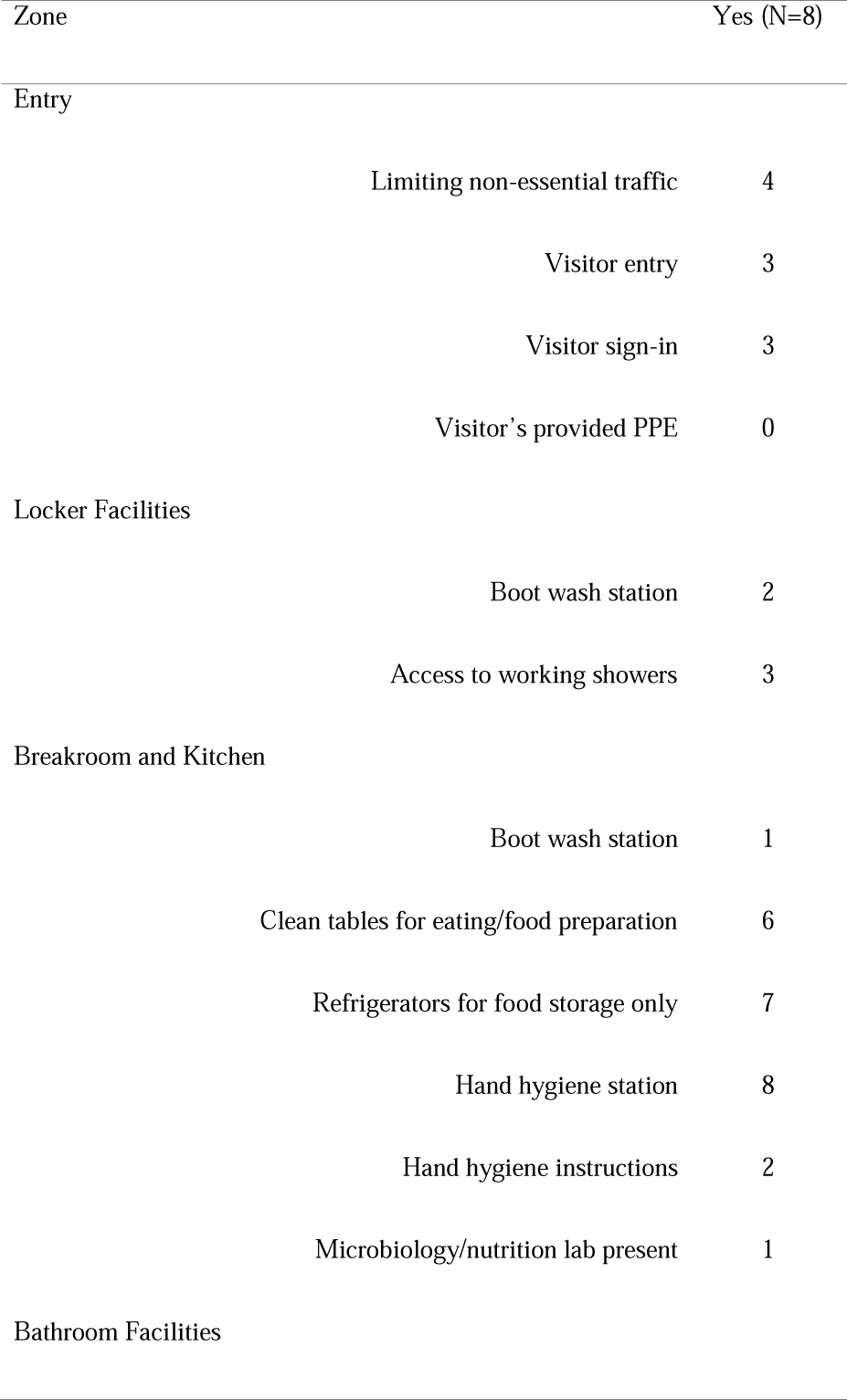
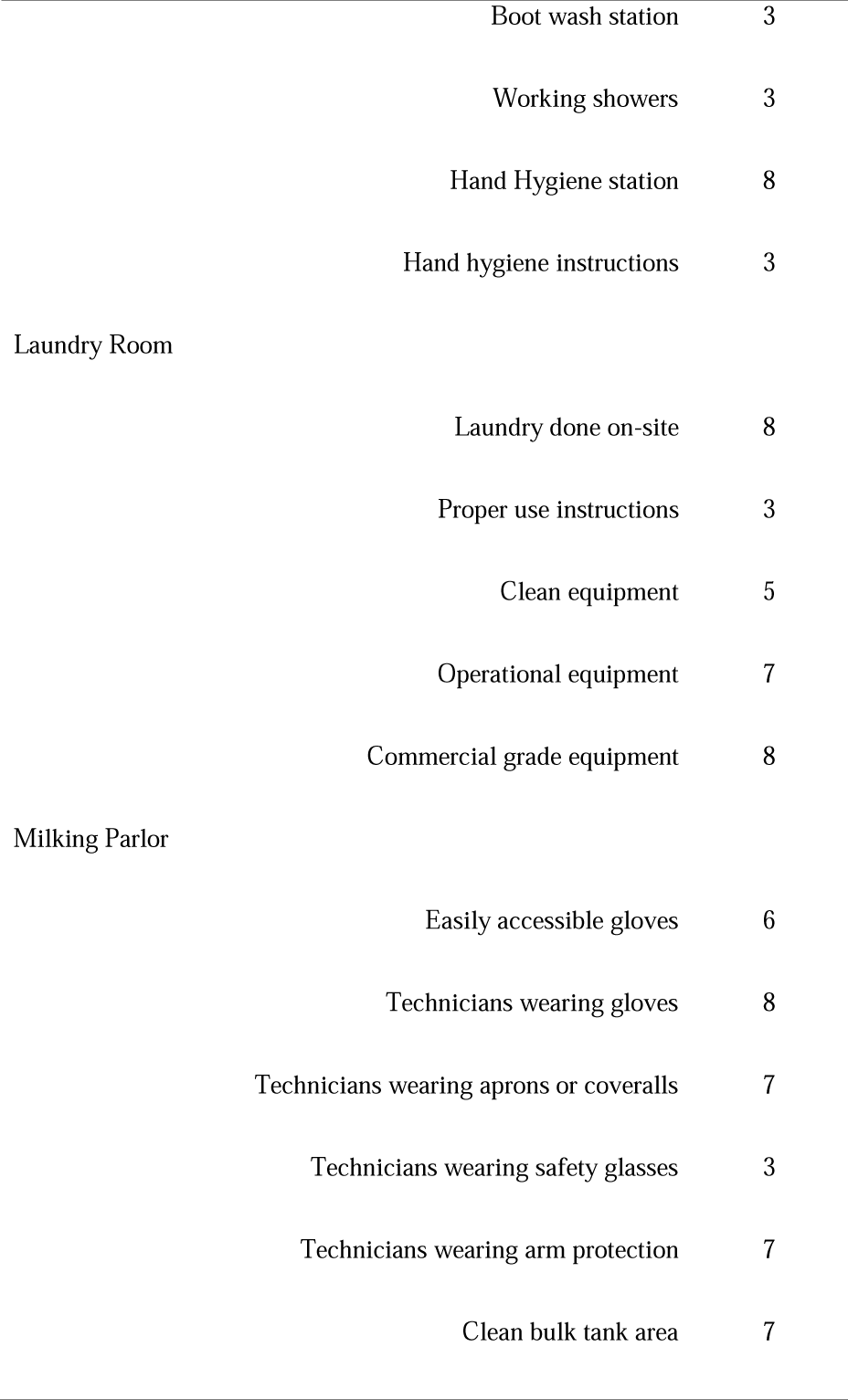
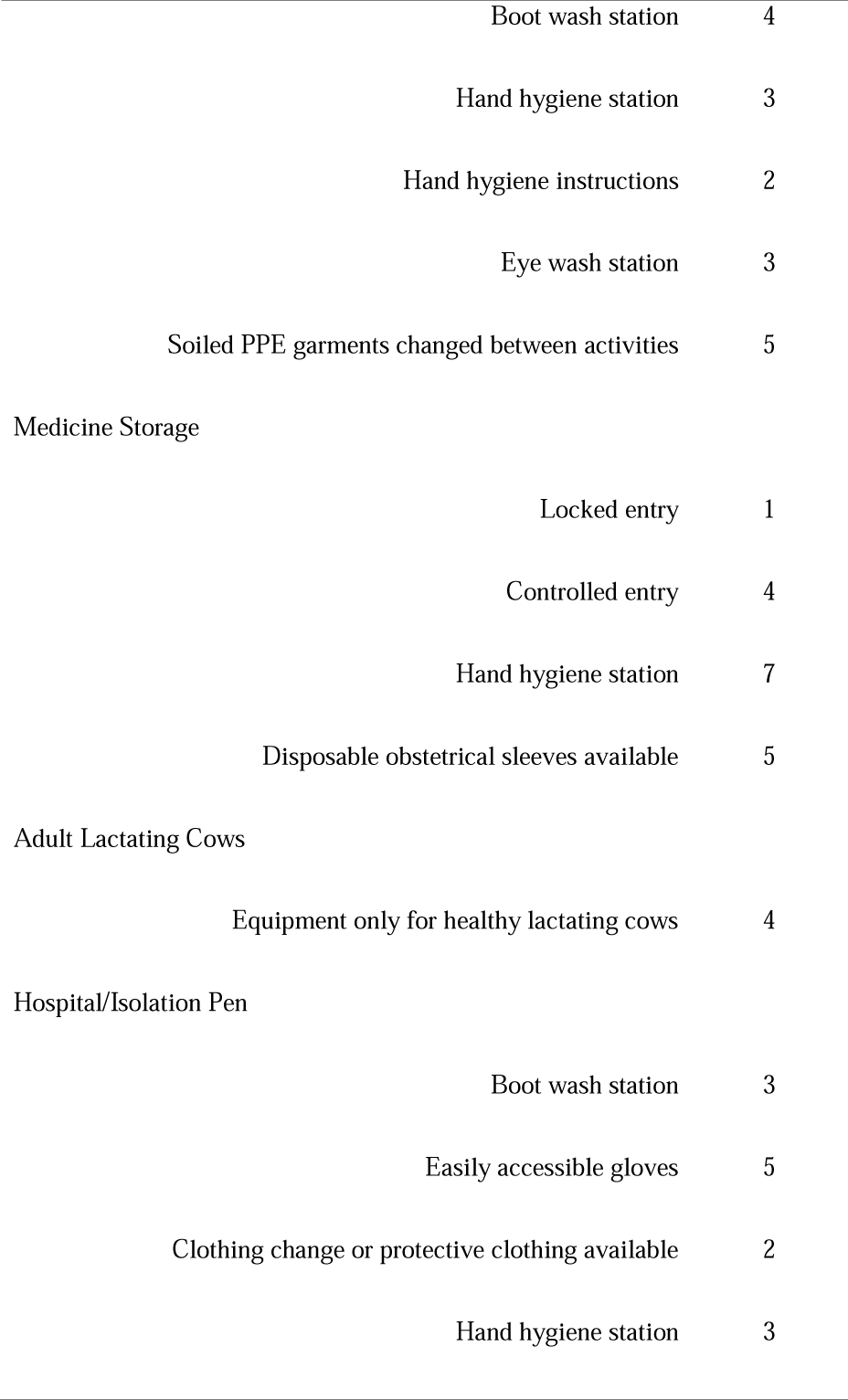
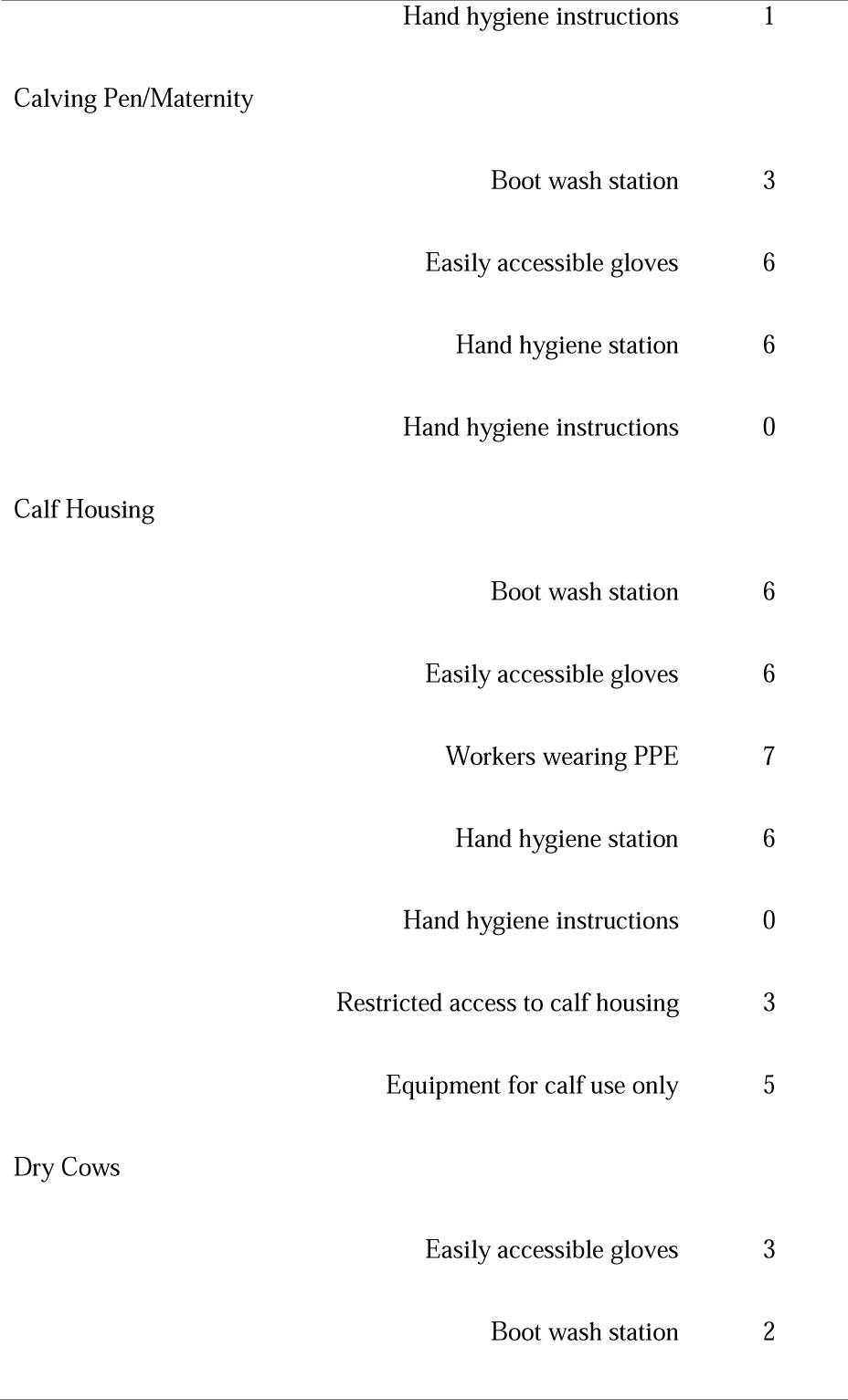
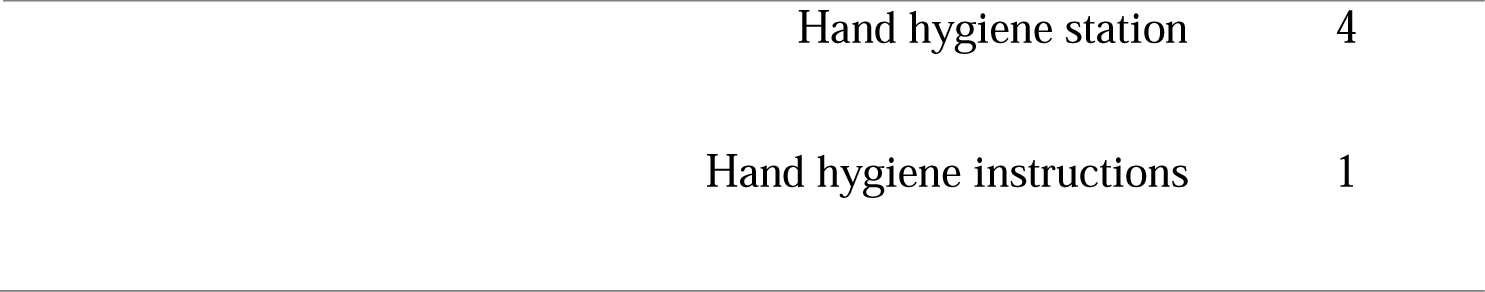
Observations of biosafety, personal protective equipment use and availability by farm zone.

| Zone | Yes (N=8) |
| --- | --- |
| Entry |  |
| Limiting non-essential traffic | 4 |
| Visitor entry | 3 |
| Visitor sign-in | 3 |
| Visitor's provided PPE | 0 |
| Locker Facilities |  |
| Boot wash station | 2 |
| Access to working showers | 3 |
| Breakroom and Kitchen |  |
| Boot wash station | 1 |
| Clean tables for eating/food preparation | 6 |
| Refrigerators for food storage only | 7 |
| Hand hygiene station | 8 |
| Hand hygiene instructions | 2 |
| Microbiology/nutrition lab present | 1 |
| Bathroom Facilities |  |

|  |  |  |
| --- | --- | --- |
|  | Boot wash station | 3 |
|  | Working showers | 3 |
|  | Hand Hygiene station | 8 |
|  | Hand hygiene instructions | 3 |
| Laundry Room |  |  |
|  | Laundry done on-site | 8 |
|  | Proper use instructions | 3 |
|  | Clean equipment | 5 |
|  | Operational equipment | 7 |
|  | Commercial grade equipment | 8 |
| Milking Parlor |  |  |
|  | Easily accessible gloves | 6 |
|  | Technicians wearing gloves | 8 |
|  | Technicians wearing aprons or coveralls | 7 |
|  | Technicians wearing safety glasses | 3 |
|  | Technicians wearing arm protection | 7 |
|  | Clean bulk tank area | 7 |

|  |  |  |
| --- | --- | --- |
|  | Boot wash station | 4 |
|  | Hand hygiene station | 3 |
|  | Hand hygiene instructions | 2 |
|  | Eye wash station | 3 |
|  | Soiled PPE garments changed between activities | 5 |
| Medicine Storage |  |  |
|  | Locked entry | 1 |
|  | Controlled entry | 4 |
|  | Hand hygiene station | 7 |
|  | Disposable obstetrical sleeves available | 5 |
| Adult Lactating Cows |  |  |
|  | Equipment only for healthy lactating cows | 4 |
| Hospital/Isolation Pen |  |  |
|  | Boot wash station | 3 |
|  | Easily accessible gloves | 5 |
|  | Clothing change or protective clothing available | 2 |
|  | Hand hygiene station | 3 |

|  |  |  |
| --- | --- | --- |
|  | Hand hygiene instructions | 1 |
| Calving Pen/Maternity |  |  |
|  | Boot wash station | 3 |
|  | Easily accessible gloves | 6 |
|  | Hand hygiene station | 6 |
|  | Hand hygiene instructions | 0 |
| Calf Housing |  |  |
|  | Boot wash station | 6 |
|  | Easily accessible gloves | 6 |
|  | Workers wearing PPE | 7 |
|  | Hand hygiene station | 6 |
|  | Hand hygiene instructions | 0 |
|  | Restricted access to calf housing | 3 |
|  | Equipment for calf use only | 5 |
| Dry Cows |  |  |
|  | Easily accessible gloves | 3 |
|  | Boot wash station | 2 |
| Hand hygiene station | 4 |
| Hand hygiene instructions | 1 |

### Environment and Physical Layout

The majority of farms had clean breakrooms (n= 6, 75%) and laundry facilities (n= 5, 62.5%) as well as locker facilities for workers to change clothes. Showers were also available on six farms (75%) (three provided showers in the locker room, three in the bathroom). On most farms, on-farm microbiology labs were kept away from human food preparation areas, although on one farm, the microbiology lab and drug storage cabinet was located in the breakroom, while on another, medication and bacterial culture supplies were also stored in close proximity to food items next to a coffee pot. Personal vehicles were not allowed past the entry on a majority of farms (n=5, 62.5%).

### Person/People

Overall, knowledge of antibiotic administration protocols was higher than anticipated and workers indicated there were only select individuals on the farm who were authorized to administer antibiotics; these workers always went to authorized workers for medications. When discussing general knowledge about antibiotics and antibiotic resistance, there was a sense workers understood the importance of good antibiotic stewardship. When discussing antimicrobial use in cattle, one worker stated:

> *“All dairy farmers have a responsibility not to overdo it*”

Similarly, there was a general understanding of the risks associated with antimicrobial resistance with two workers summarized the issue by stating:

> *“I feel like the antibiotic resistance is those can be contracted anywhere. They can start anywhere, and it comes down to how people deal with their antibiotics with the animals or with people. If we’re not prescribed the right dosage, too little or too much, we can kill it or we can just make the disease or the infections, you know, they will adapt to continue, so …”*
>
> *“I think that, well, I believe, that the bacteria, how would you say, they get used to it, they become stronger, that’s why it doesn’t work anymore*”.

While many workers demonstrated an understanding of antibiotics and resistance, this was not true for all workers. While workers may have known why it is important to use antibiotics correctly, when providing examples, they did not always appropriately identify medications as antibiotics with one worker discussing the pain reliever Tylenol when discussing experiences using antibiotics. Some workers also felt the farm managers could do more to discuss antimicrobial use and policies with the workers (Table 2).

## Discussion

We conducted focus groups and observations across eight large farms in Wisconsin to assess farm culture and behaviors potentially relating to ARG spread on farms using a systems engineering approach. Overall, farm owners/managers have successfully implemented many of the biosecurity protocols associated with mitigating ARG transmission and have implemented a positive culture around worker safety and antimicrobial use. The use of the adapted SEIPS model allowed us to identify barriers and facilitators to reducing the spread of ARGs on farms and identified several factors to considered when developing interventions to reduce the spread of ARGs.

Previous research has shown U.S. dairy farmers are not concerned about the impact of antimicrobial use on the presence of ARGs in humans and in the community (11-14). Studies have also documented lack of knowledge/belief among farmers about relationships between antimicrobial use in livestock leads and antimicrobial resistant infections in humans (12). Other research has shown most farmers do believe they are using the appropriate amount of antibiotics (15, 16) and feel they have a “moral obligation” to use antibiotics in their herds (17). A recent study of New York dairy farmers found conventional farmers had little concern for the impact of antimicrobial use on the larger community and were skeptical of policies to reduce antibiotic use on farms (11). In contrast, the workers in our study seemed to understand the importance of antimicrobial stewardship for both animal and human health. Workers in our study also demonstrated a knowledge of how wearing PPE, particularly gloves, was important for reducing transmission of ARGs. The difference in beliefs between our study and the existing literature may be due to who was interviewed. In our study, the focus was on workers while prior research focused on farm managers and owners (11, 12). Farm manager interest in antibiotic use have been reported to be associated with costs, time, and veterinary guidance (18) with conventional farmers concerned about the negative impact on animal health when reducing antimicrobial use (11). Farm workers likely have different priorities than managers and owners and further research is needed to understand these dynamics.

Workers identified substantially more facilitators to reducing ARGs than barriers. The most commonly identified facilitators were related to communication on the farm. Workers felt they had someone on the farm (herdsman, manager, or owner) they were able to talk to about any issues or needs. Most workers reported a positive culture around the use of PPE and hand hygiene. These feelings were supported by our observations on most farms. However, observations revealed boot washing and hand hygiene stations were not available in all high-risk areas and were frequently lacking in the hospital/ isolation pens. Although worker knowledge of antimicrobial resistance and its associated risks was higher than expected and previously reported, workers identified a lack of written protocols as a barrier. As previously noted, communication between workers and management was good and therefore deemed sufficient by most workers. While workers primarily identified laundry practices and access to clean clothing and coveralls as facilitators, our observer noted dirty laundry facilities on several farms. Additionally, the observer noted milk and cow towels being washed with worker clothing on several farms.

Interventions to reduce ARG spread need to be seen as both financially feasible and perceived as effective for farmers to be willing to undertake. Further education on antibiotic resistance for both the workers and managers/ owners may be a potentially effective and inexpensive intervention to reduce the spread of ARGs on farms. Workers noted a lack of education from managers/owners on this subject, though workers did have awareness of the importance of reducing resistance. Educational efforts could include signage on ways to reduce the spread of ARGs around the farms, reminders on how and when to perform hand hygiene, and when to change PPE. Furthermore, adding boot wash and hand hygiene stations may be potentially beneficial. While it may not be feasible to ask farms to add these stations where plumbing does not already exist, our observer noted sinks on several farms where no soap and/or disinfectants or towels were available.

Our study has several strengths. To our knowledge this is the first study to our knowledge to apply a systems engineering approach to assess farm workers beliefs and behaviors related to ARG transmission points. We complemented the focus group surveys with observations of worker practices on the farm as well as the availability of PPE and hand hygiene on the farms. The methods used here can be applied to future studies addressing a wide variety of farm safety topics. Additionally, these methods can be used to assess future interventions aimed at reducing ARG spread on farms.

Our study also has several limitations. Whenever workers are being observed, there is a risk they may change their behaviors while under observation (Hawthorne effect) (19) which might have led to an overestimation of glove and other PPE use. Similarly, it is possible workers told the interviewer what they thought the interviewer wanted to hear during focus group discussions To minimize these effects, observations were conducted prior to the group discussions and interviewers were trained to keep a neutral tone during discussions. Another limitation may be the size of the farms in our study – all were large operations with over 250 cows using an electronic records system to document antibiotic use, a requirement for a different aim of the project. It is possible the culture on larger farms is different than on smaller or family-run operations. We also interviewed only workers for this study and not include farm managers. As previous studies have shown, farm managers and owners may have different beliefs and priorities than workers which we were likely not captured in this study. However, farm managers’ and owners’ perceptions will be essential to developing, implementing and sustaining interventions to reduce ARG transmission

## Conclusions

Knowledge and beliefs related to ARG transmission among dairy workers were varied and viewed in a positive light, although worker knowledge was not always accurate. Interventions to reduce ARGs on dairy farms should focus on access to education on how ARGs may spread on a farm and how to reduce spread through hand hygiene practices and PPE use. The mixed methods design used (adapted SEIPS model plus direct observations) was useful in identifying barriers and facilitators relating to ARG transmission and current farm practices and identifying potential systems-level interventions. We believe this model will be of use in future studies of related issues, such as farm worker safety. Future research exploring worker and manager beliefs around ARGs is needed to better understand the extent to which knowledge and beliefs impacts ARG transmission.

## Data Availability

De-identified data is available upon reasonable request to the corresponding author.

## Acknowledgments

Funding was supplied by USDA-NIFA Food Safety Challenge Grant 2017-68003-26500. AEK was supported by a National Library of Medicine training grant to the Computation and Informatics in Biology and Medicine Training Program, USA (NLM 5T15LM007359). NS is supported grant number R01HS026226 from the Agency for Healthcare Research and Quality. The content is solely the responsibility of the authors and does not necessarily represent the official views of the Agency for Healthcare Research and Quality.

## Author Bio (first author only, unless there are only 2 authors)

Dr. Kates is a postdoctoral fellow at the University of Wisconsin School of Medicine and Public Health, Department of Medicine, Division of Infectious Disease. Her primary research interests include reducing colonization and infections with multidrug resistant pathogens as well as the role of the microbiome in human health.

**Appendix Figure 1**. Direct observations checklist.

## References

1. Prescott JF. History of Antimicrobial Usage in Agriculture: an Overview. Antimicrobial Resistance in Bacteria of Animal Origin: American Society of Microbiology; 2006.

2. Economou V, Gousia P. Agriculture and food animals as a source of antimicrobial-resistant bacteria. Infect Drug Resist. 2015;8:49–61.

3. Ruegg P, Tabone T. The relationship between antibiotic residue violations and somatic cell counts in Wisconsin dairy herds. Journal of Dairy Science. 2000;83(12):2805–9.

4. Marshall BM, Levy SB. Food animals and antimicrobials: impacts on human health. Clinical microbiology reviews. 2011;24(4):718–33.

5. Ruegg PL. Practical Food Safety Interventions for Dairy Production. Journal of Dairy Science. 2003;86:E1–E9.

6. Krueger RA, Casey MA. Focus groups: A practical guide for applied research.. Washington, DC: Sage Publications; 2014.

7. Carayon P, Schoofs Hundt A, Karsh BT, Gurses AP, Alvarado CJ, Smith M, et al. Work system design for patient safety: the SEIPS model. Qual Saf Health Care. 2006;15 Suppl 1(Suppl 1):i50–i8.

8. Musuuza JS, Hundt AS, Carayon P, Christensen K, Ngam C, Haun N, et al. Implementation of a Clostridioides difficile prevention bundle: Understanding common, unique, and conflicting work system barriers and facilitators for subprocess design. Infection control and hospital epidemiology : the official journal of the Society of Hospital Epidemiologists of America. 2019;40(8):880–8.

9. Redwood R, Knobloch MJ, Pellegrini DC, Ziegler MJ, Pulia M, Safdar N. Reducing unnecessary culturing: a systems approach to evaluating urine culture ordering and collection practices among nurses in two acute care settings. Antimicrobial resistance and infection control. 2018;7:4.

10. Leite de Campos J, Kates A, Steinberger A, Sethi A, Suen G, Shutske J, et al. Quantification of antimicrobial usage in adult cows and preweaned calves on 40 large Wisconsin dairy farms using dose-based and mass-based metrics. J Dairy Sci. 2020;Under Review.

11. Wemette M, Safi AG, Beauvais W, Ceres K, Shapiro M, Moroni P, et al. New York State dairy farmers’ perceptions of antibiotic use and resistance: A qualitative interview study. PLOS ONE. 2020;15(5):e0232937.

12. Friedman DB, Kanwat CP, Headrick ML, Patterson NJ, Neely JC, Smith LU. Importance of prudent antibiotic use on dairy farms in South Carolina: a pilot project on farmers’ knowledge, attitudes and practices. Zoonoses and public health. 2007;54(9-10):366–75.

13. Habing G, Djordjevic C, Schuenemann GM, Lakritz J. Understanding antimicrobial stewardship: Disease severity treatment thresholds and antimicrobial alternatives among organic and conventional calf producers. Preventive veterinary medicine. 2016;130:77–85.

14. Young I, Hendrick S, Parker S, Rajić A, McClure JT, Sanchez J, et al. Knowledge and attitudes towards food safety among Canadian dairy producers. Preventive veterinary medicine. 2010;94(1-2):65–76.

15. Hoe FG, Ruegg PL. Opinions and practices of wisconsin dairy producers about biosecurity and animal well-being. J Dairy Sci. 2006;89(6):2297–308.

16. Ekakoro JE, Caldwell M, Strand EB, Okafor CC. Drivers of Antimicrobial Use Practices among Tennessee Dairy Cattle Producers. Vet Med Int. 2018;2018:1836836.

17. McIntosh W, Dean W. Factors associated with the inappropriate use of antimicrobials. Zoonoses and public health. 2015;62 Suppl 1:22–8.

18. Vasquez AK, Foditsch C, Dulièpre SC, Siler JD, Just DR, Warnick LD, et al. Understanding the effect of producers’ attitudes, perceived norms, and perceived behavioral control on intentions to use antimicrobials prudently on New York dairy farms. PLoS One. 2019;14(9):e0222442.

19. Chen LF, Vander Weg MW, Hofmann DA, Reisinger HS. The Hawthorne Effect in Infection Prevention and Epidemiology. Infection Control & Hospital Epidemiology. 2015;36(12):1444–50.

