## Appendix figure 1 for "Use of a systems engineering framework to assess perceptions and practices about antimicrobial resistance of workers on large dairy farms in Wisconsin"

**Direct Observations Checklist Instrument**

Observer Name:________________________________ Date: ___ ___ / ___ ___ / ___ ___ ___ ___ Start Time: ___ ___ : ___ ___ am / pm

Farm ID: ________________________________________________________ End Time: ___ ___ : ___ ___ am / pm

| **Zone: Entry/ Parking** | ☐Not Applicable |
|---|---|
| Limiting non-essential traffic on farm[1] | ☐Yes ☐No |
| Clearly designated visitor entry | ☐Yes ☐No |
| Visitor sign-in | ☐Yes ☐No |
| Disposable PPE including boot covers (or boot wash) provided | ☐Yes ☐No |
| **Zone: Breakroom/ Kitchen** | ☐Not Applicable |
| Boot cleaning station nearby | ☐Yes ☐No |
| Refrigerators used for food storage only | ☐Yes ☐No |
| Clean tables for eating /food prep | ☐Yes ☐No |
| Sinks with soap and drying towels or blowers | ☐Yes ☐No |
| Microbiology or Nutrition lab present | ☐Yes ☐No |
| Signage reminding workers about proper hand hygiene | ☐None ☐English ☐Spanish |
| Notes: | |
| **Zone: Bathroom Facilities** | ☐Not Applicable |
| Boot wash nearby | ☐Yes ☐No |
| Shower present | ☐Yes ☐No |
| What type(s) of hand hygiene are provided:<br>☐Hand Sanitizer ☐Soap/ Water ☐Paper towels ☐ Air dryer<br>Are hand hygiene instructions provided?<br>☐English ☐Spanish ☐None | |
| Notes: | |

| **Zone: Locker Facilities.** | ☐Not Applicable |
|---|---|
| Boot wash station nearby | ☐Yes ☐No |
| Access to shower facilities with soap | ☐Yes ☐No |
| Notes: | |
| **Zone: Laundry** | ☐Not Applicable |
| Instructions on proper use | ☐None<br>☐English<br>☐Spanish |
| Instructions to use high heat and tumble dry | ☐Yes ☐No |
| Cleanliness of the washing machine<br>☐Clean ☐Mold/mildew present ☐Noticeable malodor<br>☐Visibly soiled | |
| Does the farm use an off-site service for laundry?<br>☐Yes ☐ No ☐ Partial:______________________ | |
| What is laundry room used for? | ☐Clothing<br>☐Milking Towels |
| Washing machine operational | ☐Yes ☐No |
| Dryer operational | ☐Yes ☐No |
| Are washer and/or dryer commercial/industrial or residential type | ☐Commercial<br>☐Residential |
| Where are the laundry facilities located? | |
| Notes: | |

| **Zone: Milk Room/Production** |
|---|

| **Zone: Adult Lactating Cows** | ☐Not Applicable |
|---|---|

| | |
|---|---|
| Easily accessible gloves | ☐Yes ☐No |
| Are milking technicians wearing gloves | ☐Yes ☐No |
| Are milking technicians wearing plastic aprons | ☐Yes ☐No |
| Are milking technicians wearing coveralls | ☐Yes ☐No |
| Are milking technicians wearing safety glasses | ☐Yes ☐No |
| Do milking technicians have arm protection | ☐Yes ☐No |
| Cleanliness of bulk tank area<br>Signage designating clean area<br>☐ Yes ☐No | ☐Clean<br>☐Some soiling<br>☐Visibly soiled |
| Eye wash station | ☐Yes ☐No |
| Are soiled PPE garments changed prior to changing milkhouse activities | ☐Yes ☐No |
| Cloth towel or disposable wipes? | ☐Cloth towels<br>☐Disposable wipes |

What type(s) of hand hygiene are provided:
☐Hand Sanitizer ☐Soap/ Water ☐Paper towels ☐ Air dryer
Are hand hygiene instructions provided?
☐English ☐Spanish ☐None

| | |
|---|---|
| Boot cleaning station nearby | ☐Yes ☐No |

Notes on milking production:

**Zone: Medicine Storage** ☐Not Applicable

| | |
|---|---|
| Locked entry | ☐Yes ☐No |
| Controlled access | ☐Yes ☐No |
| Gloves/hand hygiene present | ☐Yes ☐No |
| Plastic/disposable obstetrical sleeves available | ☐Yes ☐No |

Notes:

| | |
|---|---|
| Storage for equipment used only around healthy lactating cows | ☐Yes ☐No |

Notes:

**Zone: Sick Cow Pen/ Hospital/ Isolation** ☐Not Applicable

| | |
|---|---|
| Boot wash station or disposable boot covers | ☐Yes ☐No |
| Easily accessible gloves | ☐Yes ☐No |

Are workers either able to change their clothing or provided with protective clothing?
☐Yes, changing facility ☐Yes, coveralls provided or plastic aprons
☐No ☐Other:____________________________________________

Are workers wearing PPE?
☐Gloves ☐Boot Covers ☐Coveralls or Aprons ☐No ☐N/A
☐Other: ____________________________________________

What type(s) of hand hygiene are provided:
☐Hand Sanitizer ☐Soap/ Water ☐Paper towels ☐ Air dryer ☐No
Are hand hygiene instructions provided?
☐English ☐Spanish ☐None

Notes:

**Zone: Calving Pen/Maternity** (Cont. next page) ☐Not Applicable

| | |
|---|---|
| Boot wash station nearby | ☐Yes ☐No |
| Easily accessible gloves | ☐Yes ☐No |
| Access to hand hygiene | ☐Yes ☐No |
| Are workers wearing PPE | ☐Yes ☐No |

What type(s) of hand hygiene are provided:
☐Hand Sanitizer ☐Soap and Water ☐Paper towels
☐ Air dryer

**Zone: Calving Pen/ Maternity** (cont.) ☐Not Applicable

**Zone: Cull Cows and Trucks** ☐Not Applicable

Are hand hygiene instructions provided?
☐English ☐Spanish ☐None

Are workers wearing PPE?
☐Gloves ☐Boot Covers ☐Coveralls or Aprons ☐No ☐N/A

☐Other: ______________________________

Notes:

**Zone: Baby Calf Hutches/ Housing** ☐Not Applicable

| | |
|---|---|
| Boot wash station or disposable boot covers | ☐Yes ☐No |
| Easily accessible gloves | ☐Yes ☐No |

Are workers wearing PPE?
☐Gloves ☐Boots or Covers ☐Coveralls or Aprons ☐No

☐Other: ______________________________

What type(s) of hand hygiene are provided:
☐Hand Sanitizer ☐Soap/ Water ☐Paper towels ☐ Air dryer

Are hand hygiene instructions provided?
☐English ☐Spanish ☐None

| | |
|---|---|
| Restricted access to calf housing? | ☐Yes ☐No |
| Equipment storage only for calves (e.g. shovels, buckets, halters, etc.) | ☐Yes ☐No |

Notes:

How are carcasses disposed of?
☐Disposal on site ☐Licensed Collection Company
**If on site**: ☐Burial ☐Incineration ☐Composting ☐Rendering

Protective clothing provided
☐Coveralls ☐Aprons ☐Gloves ☐Boot covers ☐None ☐NA

| | |
|---|---|
| Are workers required to change clothing prior to handling other animals | ☐Yes ☐No |
| Are there signs limiting who and what vehicles can be in the zone? | ☐Yes ☐No |
| Is there a place to clean trucks and tires | ☐Yes ☐No |

What type(s) of hand hygiene are provided:
☐Hand Sanitizer ☐Soap/ Water ☐Paper towels ☐ Air dryer

Are hand hygiene instructions provided?
☐English ☐Spanish ☐None

Notes:

**Zone: Dry Cows** ☐Not Applicable

| | |
|---|---|
| Are gloves easily accessible | ☐Yes ☐No |
| Boot wash station or disposable boot covers | ☐Yes ☐No |
| Are workers using gloves | ☐Yes ☐No |

What type(s) of hand hygiene are nearby:
☐Hand Sanitizer ☐Soap/ Water ☐Paper towels ☐ Air dryer

Are hand hygiene instructions provided?
☐English ☐Spanish ☐Both

Notes

| **General Biosecurity Observations:** | | **Notes:** |
|---|---|---|
| Do workers change or launder clothes or shoes prior to leaving the farm? | ☐Yes ☐No | |

| | |
|---|---|
| Are coveralls provided for workers? | ☐Yes ☐No |
| Are boots or shoe coverings provided for workers? | ☐Yes ☐No |
| Where do workers typically eat? | |
| Are personal vehicles able or allowed to be driven where animals may walk or be transported? | ☐Yes ☐No |
| How often are employees encouraged to change PPE? | |
| Obstetrical chains? | |
| Hot water heater heats to temp and supplies sufficient hot water | |

Which of the following disinfectants, if any, did you see on the farm?

☐Bleach/ chlorine based (Clorox) ☐Chlorhexidine (Nolvasan) ☐ Iodophors (Betadine/ Weladol) ☐Oxidizers (Virkon/ Oxy-Sept 333) ☐Ammonium (Roccal D Plus) ☐ Phenolic (Pine-Sol, One Stroke, Osyl) ☐No ☐Other:

### Other Observations
